# Genome-Wide Association Study of Genetic Variants Associated with Lower Extremity Amputation Risk in Peripheral Artery Disease

**DOI:** 10.64898/2025.12.17.25342479

**Authors:** Rajashekar Korutla, Tanisha Garg, Michael P. Wilczek, Elsie G. Ross, Saeed Amal

**Affiliations:** The Roux Institute of Northeastern University, Portland, ME 04101, USA; Department of Bioengineering, College of Engineering, Northeastern University, Boston, MA 02120, USA; Department of Surgery, Division of Vascular Surgery, University of California, San Diego School of Medicine, La Jolla, San Diego, CA 92037, USA; College of Science, The Roux Institute, Northeastern University, Portland, ME 04101, USA

**Keywords:** Peripheral artery disease, amputation, genome-wide association study, genetic risk factors, single nucleotide polymorphisms, Firth correction

## Abstract

Peripheral artery disease (PAD) is a global health burden affecting over 200 million individuals and is frequently complicated by limb-threatening ischemia, leading to major amputations. Despite known clinical risk factors, the genetic basis underlying amputation risk in PAD remains poorly defined. In this study, we performed a multi-pronged genome-wide association study (GWAS) to identify genetic variants associated with lower extremity amputation in patients with PAD, using data from the All of Us Research Program. Two analytical strategies were employed: a targeted GWAS using ClinVar variants on the full cohort and a comprehensive genome-wide association study using Allele Count/Allele Frequency (ACAF) data on a balanced subset. The ClinVar analysis of 118,871 variants in 14,771 PAD patients (613 with amputation, 14,158 without) identified 3 suggestive associations with a genomic inflation factor of 1.046. The ACAF analysis of 7,784,837 quality-controlled variants in 804 balanced samples (399 cases, 405 controls) yielded 35 suggestive associations (p < 1×10□□) with a genomic inflation factor of 1.017. No variants achieved suggestive significance in both analyses. These findings highlight candidate loci for further validation and may inform future development of risk prediction tools and targeted interventions to reduce limb loss in PAD.

## Introduction

Peripheral artery disease (PAD) represents a significant global health burden, affecting over 200 million individuals worldwide and increasing in prevalence with age (Fowkes et al., 2013; Leeper et al., 2012). PAD is characterized by atherosclerotic narrowing of the peripheral arteries, predominantly in the lower extremities, leading to reduced blood flow and tissue ischemia (Norgren et al., 2007; Morley et al., 2018). According to the 2017 ESC Guidelines, PAD is a progressive, systemic condition that requires comprehensive diagnostic and therapeutic strategies to reduce cardiovascular risk and prevent limb-threatening complications (Ricco et al., 2017). **S**imilarly, the 2016 AHA/ACC guidelines emphasize early detection and comprehensive management of lower extremity PAD to reduce both cardiovascular and limb-related morbidity (Gerhard-Herman et al., 2016).

While many PAD patients remain asymptomatic or present with intermittent claudication, a subset progresses to critical limb ischemia (CLI), the most severe manifestation of PAD, carrying a high risk of limb loss (Conte et al., 2019). Amputation represents a devastating complication of PAD that significantly impacts patient quality of life, functional status, and long-term survival (Mustapha et al., 2018). Despite advances in revascularization techniques and medical therapy, amputation rates remain substantial, particularly among certain demographic groups and those with comorbidities such as diabetes mellitus (Goodney et al., 2015). Diabetes is a major risk factor for both the development and progression of PAD and significantly increases the likelihood of limb loss due to impaired vascular and metabolic responses (Fox et al., 2015, korutla et al., 2025). The 5-year mortality rate following major amputation exceeds 50%, highlighting the severe implications of this outcome (Jones et al., 2014). Moreover, patients with PAD often have polyvascular disease, which has been shown to significantly increase the risk of major cardiovascular events and may compound the risk of limb loss (Gutierrez et al., 2018).

While clinical risk factors for PAD progression and amputation have been extensively studied, the genetic determinants underlying amputation risk remain poorly understood. Previous genetic studies in PAD have identified several loci associated with disease susceptibility and progression (Matsukura et al., 2015; Wassel et al., 2012), but specific genetic variants predisposing to amputation have not been thoroughly investigated.

Understanding the genetic architecture of amputation risk in PAD could potentially lead to improved risk stratification, personalized preventive strategies, and novel therapeutic targets. Despite advances in understanding PAD biology, the genetic underpinnings of disease susceptibility and progression remain incompletely defined, particularly for severe outcomes such as amputation (Kullo & Leeper, 2015).

Genome-wide association studies (GWAS) have emerged as a powerful approach for identifying genetic variants associated with complex traits and diseases (Tam et al., 2019). This methodology has successfully uncovered numerous susceptibility loci for cardiovascular diseases, including coronary artery disease, stroke, and peripheral artery disease (“A Comprehensive 1000 Genomes–based Genome-wide Association Meta-analysis of coronary artery disease,” 2015; Malik et al., 2018; Klarin et al., 2019). However, to our knowledge, no GWAS has specifically investigated genetic determinants of amputation risk in PAD patients.

In this study, we conducted a comprehensive GWAS analysis to identify genetic variants associated with amputation in PAD patients. We employed multiple methodological approaches, including targeted analysis using the clinically relevant ClinVar dataset on the full cohort and a genome-wide analysis using the Allele Count/Allele Frequency (ACAF) dataset on a balanced subset.

## Methods

### Study Population

The study cohort consisted of patients with confirmed PAD from the *All of Us* Research Program. We identified PAD patients using established clinical criteria and diagnostic codes. From this cohort, we further classified patients into two groups: those who had undergone amputation (cases) and those who had not (controls). Patient data were extracted from electronic health records and linked to genetic information. This study was conducted in accordance with relevant ethical guidelines.

### Patient Cohorts

For the ClinVar analysis, we utilized the full cohort of 14,771 PAD patients identified in the All of Us database, comprising 613 patients who had undergone amputation and 14,158 who had not. Demographic data, including age and sex, were available for 9,607 patients (65% of the cohort). After filtering for genetic data availability and complete covariate information, 7,558 patients were included in the final analysis.

For the ACAF analysis, we employed a balanced case-control design to optimize statistical power and computational efficiency. From the initial 14,771 PAD patients (613 with amputation [4.2%], 14,158 without amputation [95.8%]), we observed significant demographic differences between groups. Amputated patients were younger (median age 60 vs 68 years), more likely to be male (63.4% vs 50.2%), Black/African American (25.8% vs 16.0%), or Hispanic/Latino (18.4% vs 11.4%). To create a balanced cohort, we performed optimal one to one matching using the Hungarian algorithm (linear sum assignment) based on Euclidean distances calculated from normalized features including age, race, ethnicity, sex, number of observations, and length of observation. Features were standardized using StandardScaler, and categorical variables were label-encoded. This ensured each non-amputation patient was matched to exactly one amputation patient, minimizing the overall matching distance. The matching quality was excellent with a mean distance of 0.29 (median: 0.21, max: 1.59). We verified that no patients appeared in both groups.

From the 534 matched pairs, only 859 patients (5.8% of the original cohort) had whole genome sequencing data available in the ACAF dataset: 424 amputation cases and 435 controls. After removing related individuals, the final analysis cohort consisted of 804 individuals: 399 amputation cases and 405 controls.

### Genotyping and Quality Control

Genetic data were obtained from whole-genome sequencing performed as part of the *All of Us* Research Program.

### ClinVar Analysis

For the targeted analysis, we utilized the pre-processed ClinVar dataset, focusing on clinically relevant variants. Quality control procedures included filters for minor allele frequency (MAF > 0.01), Hardy-Weinberg equilibrium (p > 1×10□¹□), and call rate (> 0.95). The initial ClinVar dataset contained 142,503 variants. After quality control, 118,871 variants remained for analysis.

### ACAF Analysis

For the comprehensive genome-wide analysis, we employed the ACAF dataset version 8. Quality control procedures included:

- Call rate filter (> 0.95): Removed 31,671,940 variants
- Minor allele frequency filter (> 0.05): Removed 76,514,080 variants
- Hardy-Weinberg equilibrium filter (p > 1×10□□): Removed 485,562 variants
- Monomorphic variant check: Removed 0 variants

Starting with 116,456,419 variants, 7,784,837 variants remained after quality control. We removed 55 related sample pairs, resulting in 804 individuals for analysis.

### Statistical Analysis

We conducted two separate GWAS analyses with different designs based on data characteristics and computational constraints:

### ClinVar Analysis

Logistic regression was performed on 118,871 quality-controlled variants using Wald tests. The initial cohort of 14,771 PAD patients was reduced to 7,558 samples (51%) with complete data for all covariates after filtering for genetic data availability, demographic completeness, and relatedness. Covariates included age, sex, and the first three principal components of ancestry. The analysis utilized the natural case-control distribution (613:14,158) given the focused nature of testing pre-selected clinically relevant variants.

### ACAF Analysis

A comprehensive genome-wide analysis was performed on 7,784,837 quality-controlled variants using logistic regression with Wald tests. Initial attempts to include age and sex as covariates resulted in convergence failures ("exploded at Newton iteration 1" errors). After systematic optimization, the final model included only the first three principal components of ancestry as covariates, achieving successful convergence across all variants. The balanced design (399:405) was specifically chosen to maximize statistical power and ensure computational stability when testing millions of variants.

For both analyses:

- Statistical significance was defined as p < 5×10□□ for genome-wide significance and p < 1×10□□ for suggestive associations.
- Genomic inflation factor (λ) was calculated to assess potential population stratification or systematic biases.
- Results were visualized using Manhattan plots and quantile-quantile (QQ) plots.

### Bioinformatic Analysis

#### Variant Annotation and Functional Prediction

All 38 suggestive variants (p < 1×10□□) from both ClinVar and ACAF analyses underwent comprehensive functional annotation using multiple bioinformatics resources.

### Variant Effect Prediction

We annotated variants using the Ensembl Variant Effect Predictor (VEP) REST API (GRCh38/hg38) to determine molecular consequences, affected genes, and gene biotypes. For intergenic variants, we identified the nearest gene within a 500kb window using Ensembl’s overlap API. Complex variants such as deletions required alternative API query formats using dash notation for successful annotation retrieval.

### Pathogenicity Assessment

We evaluated variant deleteriousness using the Combined Annotation Dependent Depletion (CADD) framework version 1.7. Both raw scores and PHRED-scaled scores were obtained, where PHRED scores of 10, 20, and 30 indicate variants in the top 10%, 1%, and 0.1% of deleterious variants, respectively.

### Population Frequency Analysis

Allele frequencies across diverse populations were obtained from the Genome Aggregation Database (gnomAD v4) via GraphQL API queries. Maximum allele frequencies were calculated across both exome and genome datasets for comprehensive coverage. Population-specific frequencies were assessed for European (EUR), African (AFR), East Asian (EAS), South Asian (SAS), and Latino/Admixed American (AMR) populations.

#### Expression and Regulatory Analysis

### Expression Quantitative Trait Loci (eQTL) Analysis

We queried the GTEx Portal v10 to identify whether variants act as expression or splicing quantitative trait loci. Each variant was searched by rsID to identify cis-eQTL effects (within 1 Mb of target genes) and splicing QTL effects across 54 tissues, with emphasis on vascular tissues (tibial artery, aorta), adipose tissue, and blood.

### Regulatory Element Analysis

We employed two complementary approaches to identify regulatory variants. First, all variants were queried using RegulomeDB v2.2, which integrates data from ENCODE, Roadmap Epigenomics, and other consortia, assigning categorical scores from 1a-7 based on regulatory evidence strength. Second, variants with the strongest RegulomeDB scores were validated using ENCODE SCREEN v3 to identify candidate cis-regulatory elements based on chromatin accessibility and histone modifications across 1,518 biosamples.

### Chromatin Accessibility Assessment

We queried the ENCODE database REST API for DNase-seq and ATAC-seq experiments, prioritizing vascular-relevant cell types including endothelial cells, vascular smooth muscle cells, and monocytes. Regulatory features within ±5kb of each variant were identified using the Ensembl Regulation database.

### Statistical Refinement Analyses

#### Conditional Analysis

To identify independent signals within associated loci, we performed stepwise conditional analysis on seven genomic regions containing multiple suggestive variants. Genotype data were extracted for expanded windows (100-200 kb) around each cluster, and association testing was repeated while adjusting for the lead variant genotype dosage and the same covariates used in the primary analysis.

### Alternative Control Analysis

We evaluated the impact of control selection by comparing our extreme phenotype design (1:1 matched cases and controls) against a traditional approach using all available PAD patients without amputation as controls (1:17.9 ratio). Association statistics were compared between approaches for all 38 suggestive variants.

### Covariate Adjustment Analysis

To assess robustness to demographic confounding, we compared models with increasing covariate complexity: Model 1 (baseline) with only ancestry principal components, and Model 2 with principal components plus age at enrollment. Additional models incorporating sex failed to converge due to numerical instability.

#### Regional Visualization

For the six most significant loci, we generated regional association plots spanning 1-2 Mb windows centered on lead variants. Plots displayed -log□□(p-values) against chromosomal position, with variants colored by physical distance from the lead SNP as a proxy for linkage disequilibrium: purple diamond (lead variant), orange (<10 kb), gold (10-50 kb), light green (50-100 kb), and sky blue (>100 kb). Horizontal reference lines indicated genome-wide significance (p = 5×10□□) and suggestive significance (p = 1×10□□) thresholds. Gene annotations were extracted and filtered to display relevant protein-coding genes and long non-coding RNAs within each region.

## Results

### ClinVar Analysis Results

The targeted GWAS using the ClinVar dataset did not identify any variants reaching genome-wide significance (p < 5×10□□). However, three variants showed suggestive associations (p < 1×10□□) as shown in the table 1.

**Table 1:** 3 Suggestive SNPs from Clinvar analysis.

| Chr:Position | rsID | Ref>Alt | Beta | P-value | OR | Gene/Nearest Gene | Consequence | Distance (bp) | MAF (%) | eQTL | CADD PHRED |
| --- | --- | --- | --- | --- | --- | --- | --- | --- | --- | --- | --- |
| chr6:129 | rs6242 | G>A | 0.88 | 1.00x1 | 2.41 | LAMA | intron_ | 0 | 2.15 | Yes | 0.4 |
| 165442 | 0977 |  |  | 0 □ □ |  | 2 | variant |  |  |  |  |
| chr22:50<br>076844 | rs6010<br>165 | G>A | 0.484 | 2.00×1<br>0 □ □ | 1.62 | MLC1 | synony<br>mous_<br>variant | 0 | 11.4 | Yes | 0.54 |
| chr22:50<br>076841 | rs2676<br>07236 | T>C | 0.484 | 2.00×1<br>0 □ □ | 1.62 | MLC1 | splice_<br>region_<br>_varia<br>nt | 0 | 10.8 | No | 0.02 |

The genomic inflation factor (λ) was 1.046.

The Manhattan plot in Figure 1 shows peaks of suggestive significance, particularly on chromosomes 6 and 22, while the QQ plot in Figure 2 showed some deviation from the expected distribution at the tail end, suggesting potential associations.

**Figure 1:**
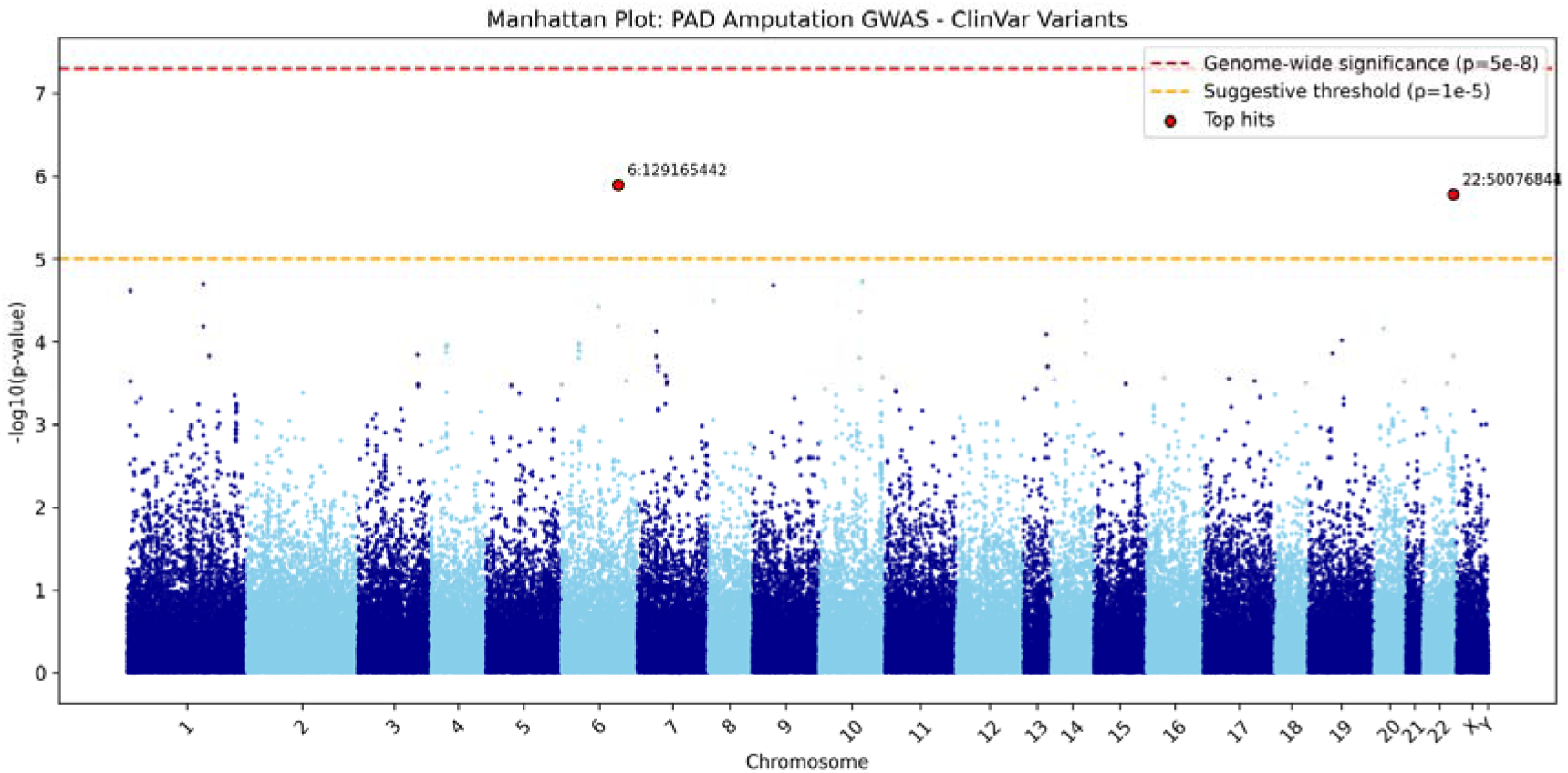
Manhattan Plot on Clinvar Analysis

**Figure 2:**
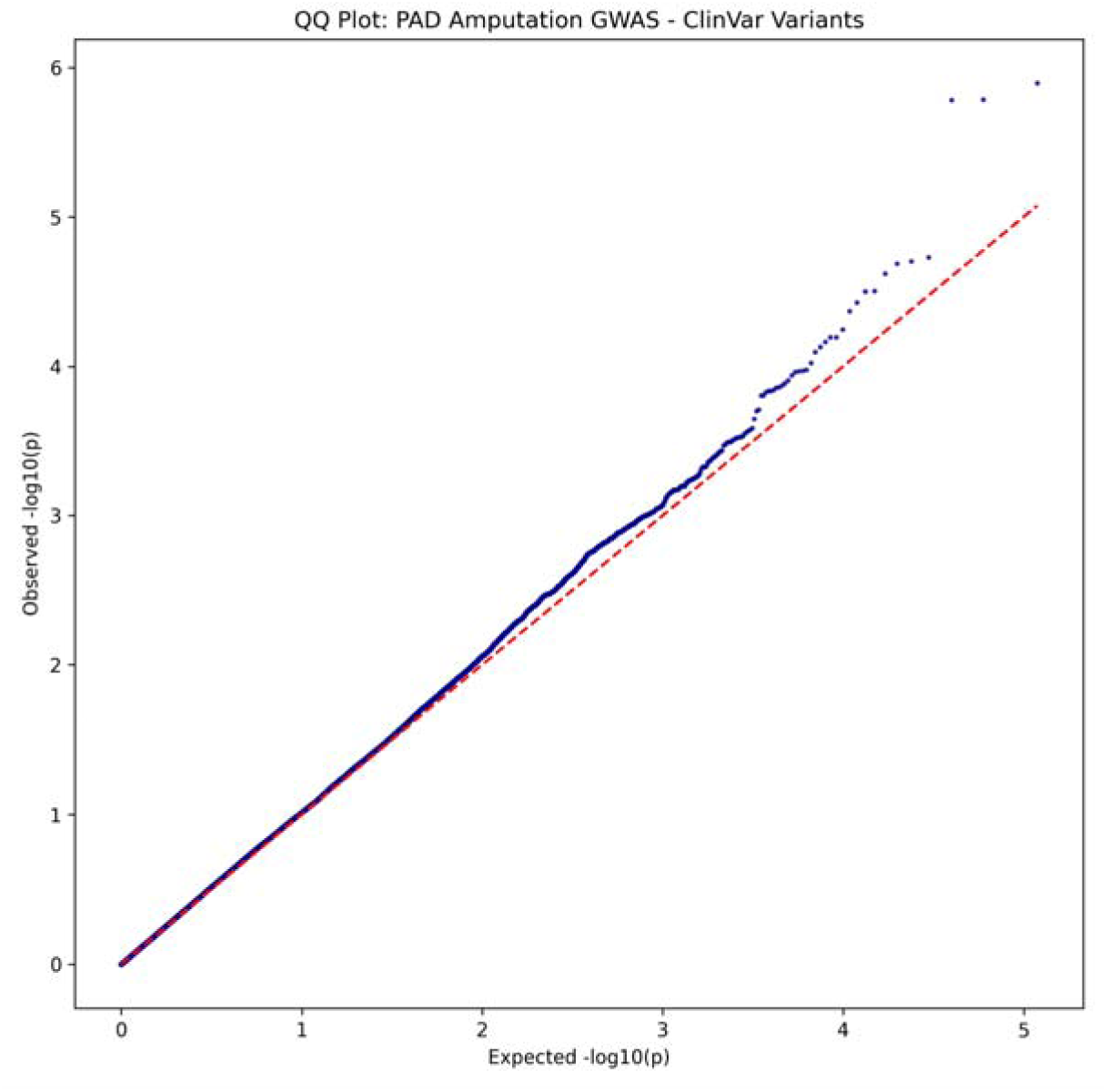
QQ Plot on Clinvar Analysis

### ACAF Analysis Results

The comprehensive GWAS using the ACAF dataset tested 7,784,837 variants in 804 samples. No variants reached genome-wide significance. 35 variants showed suggestive associations (p < 1×10□□). The genomic inflation factor (λ) was 1.017. The suggestive variants can be seen in Table 2.

**Figure 3:**
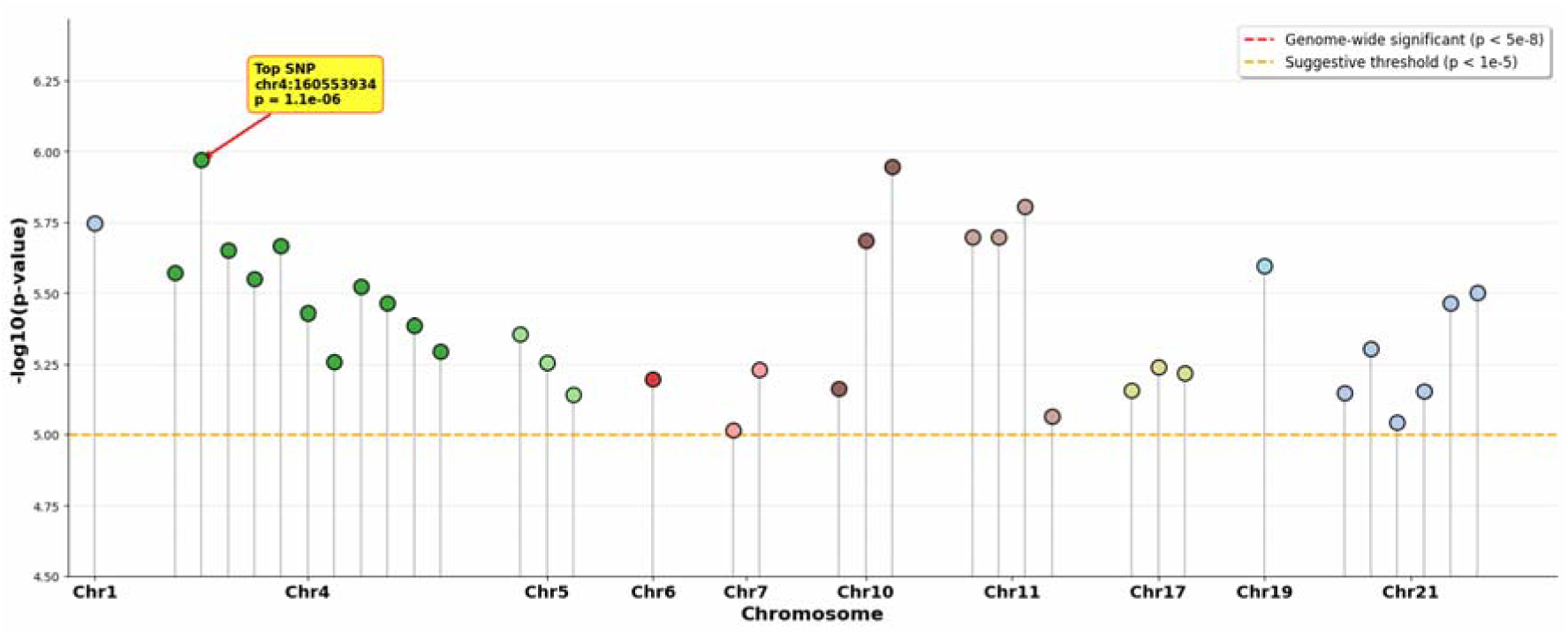
Manhattan Plot of ACAF analysis

**Figure 4:**
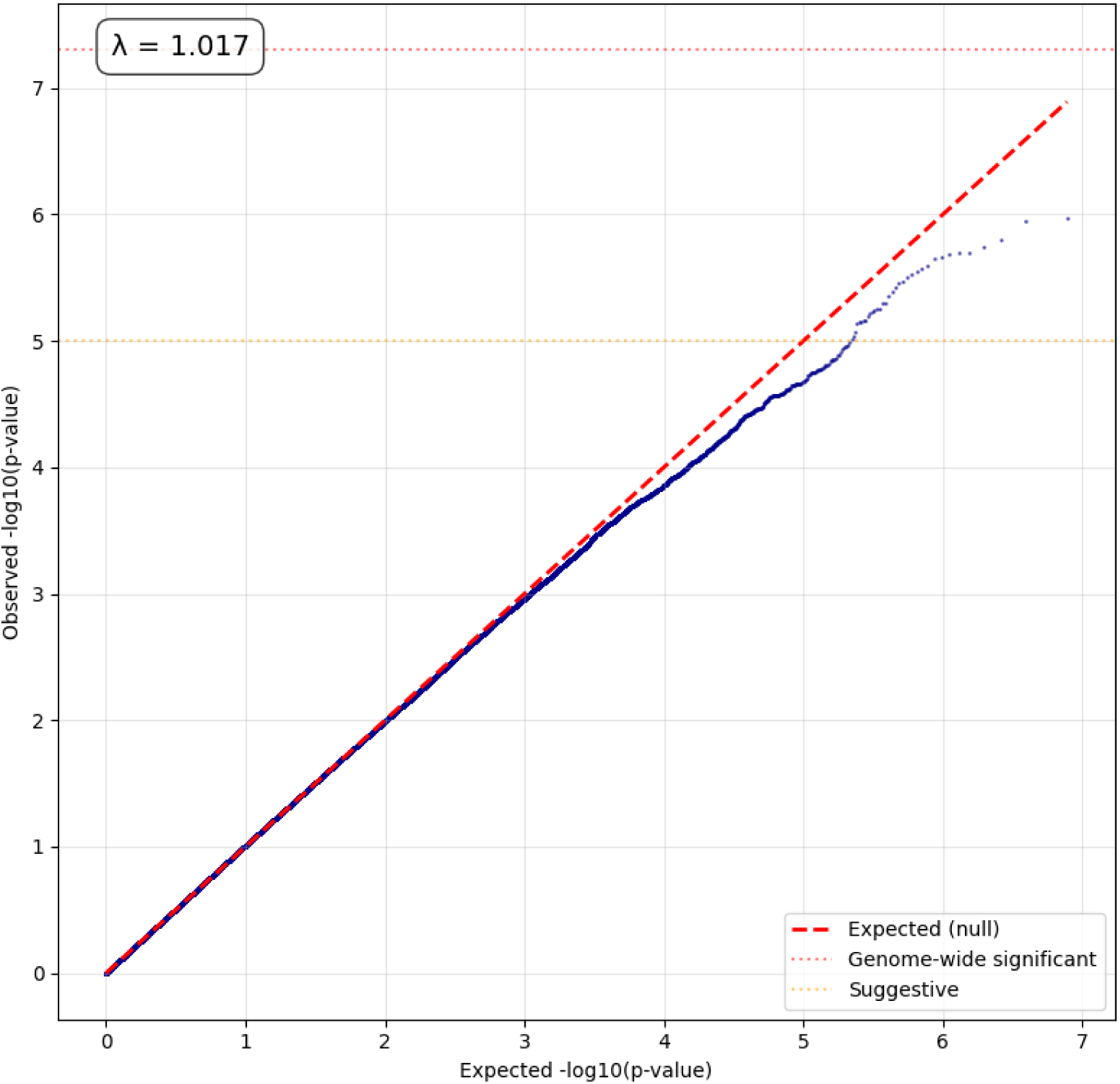
QQ Plot of ACAF analysis

**Table 2:**
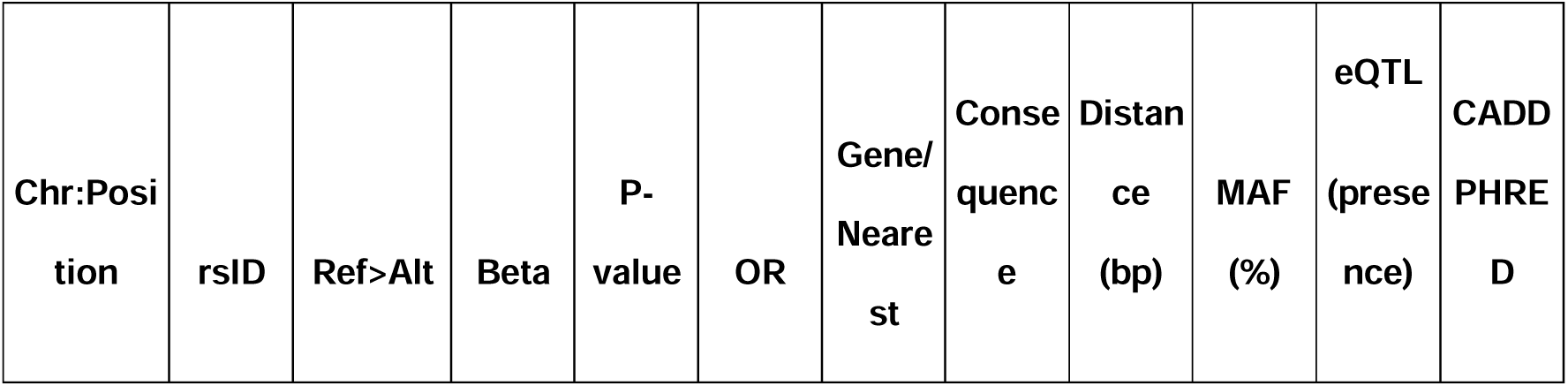
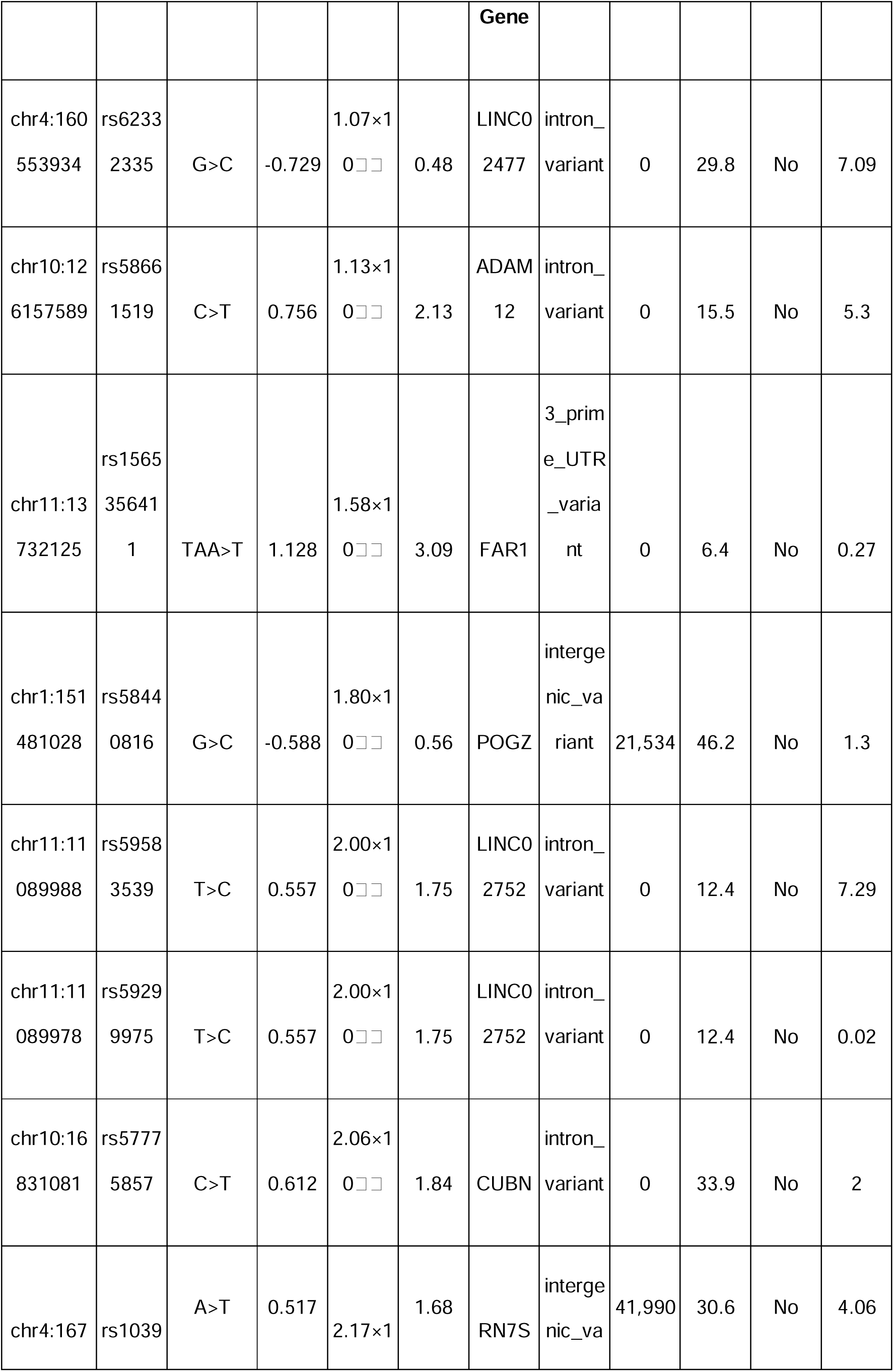
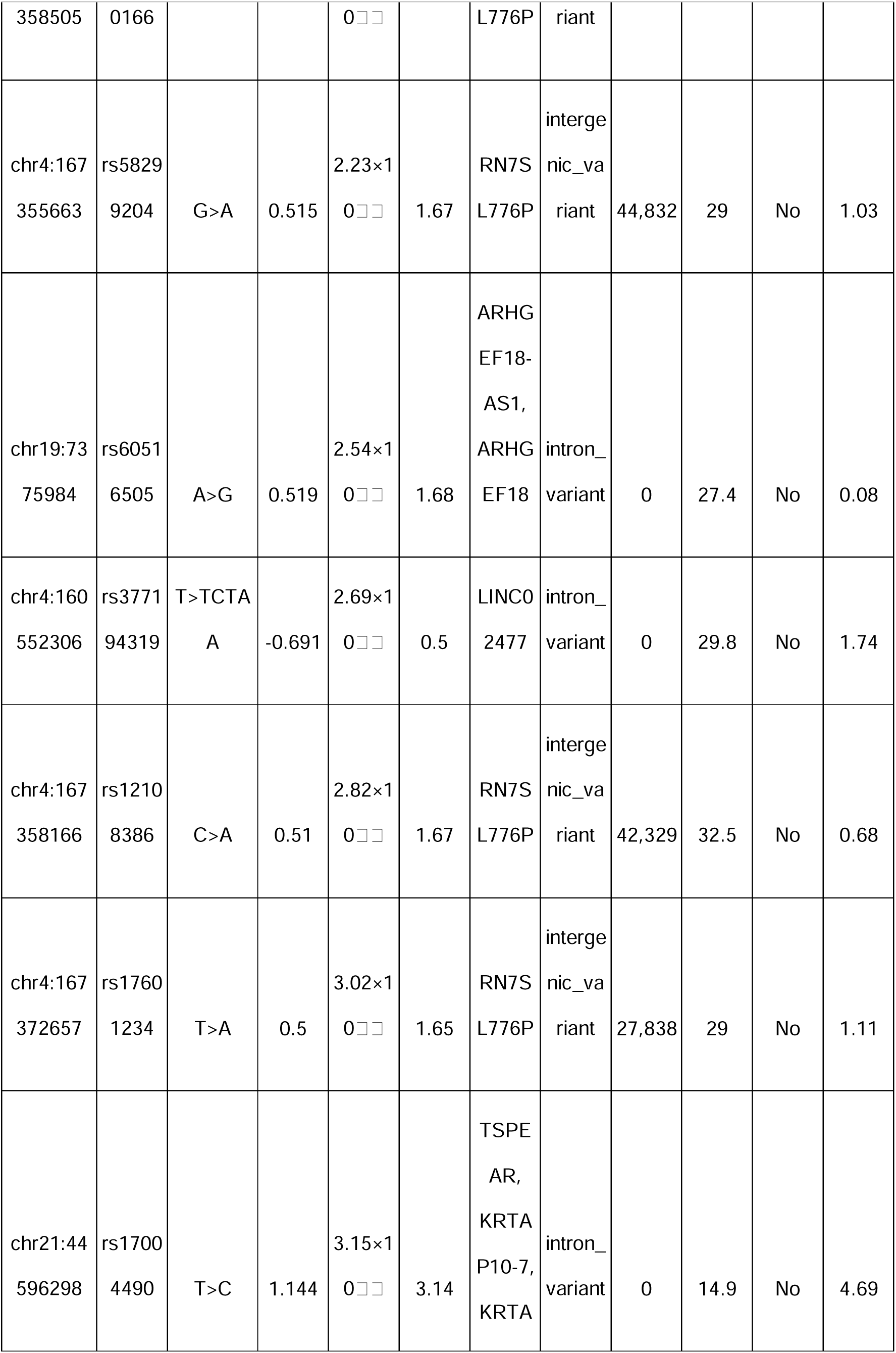
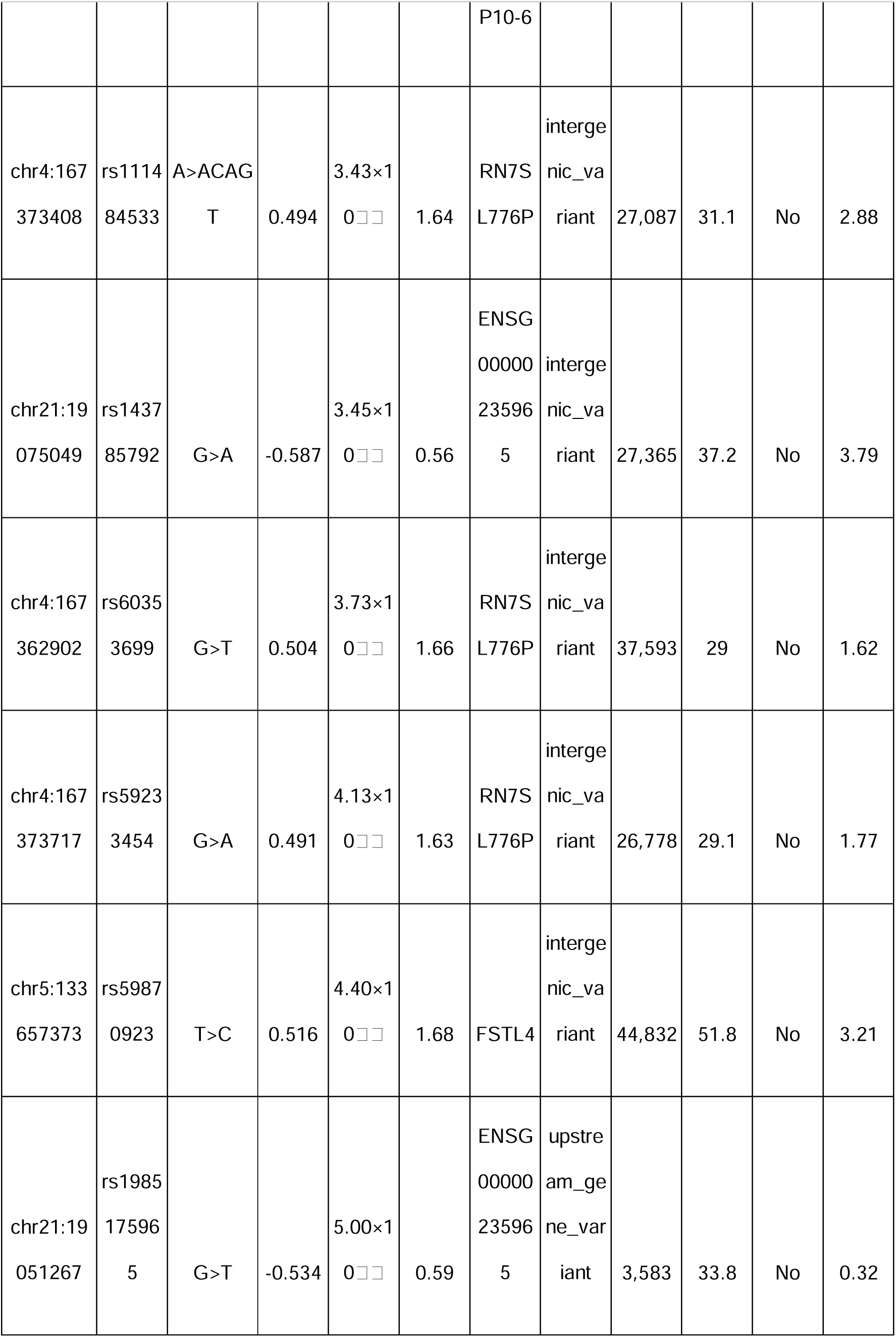
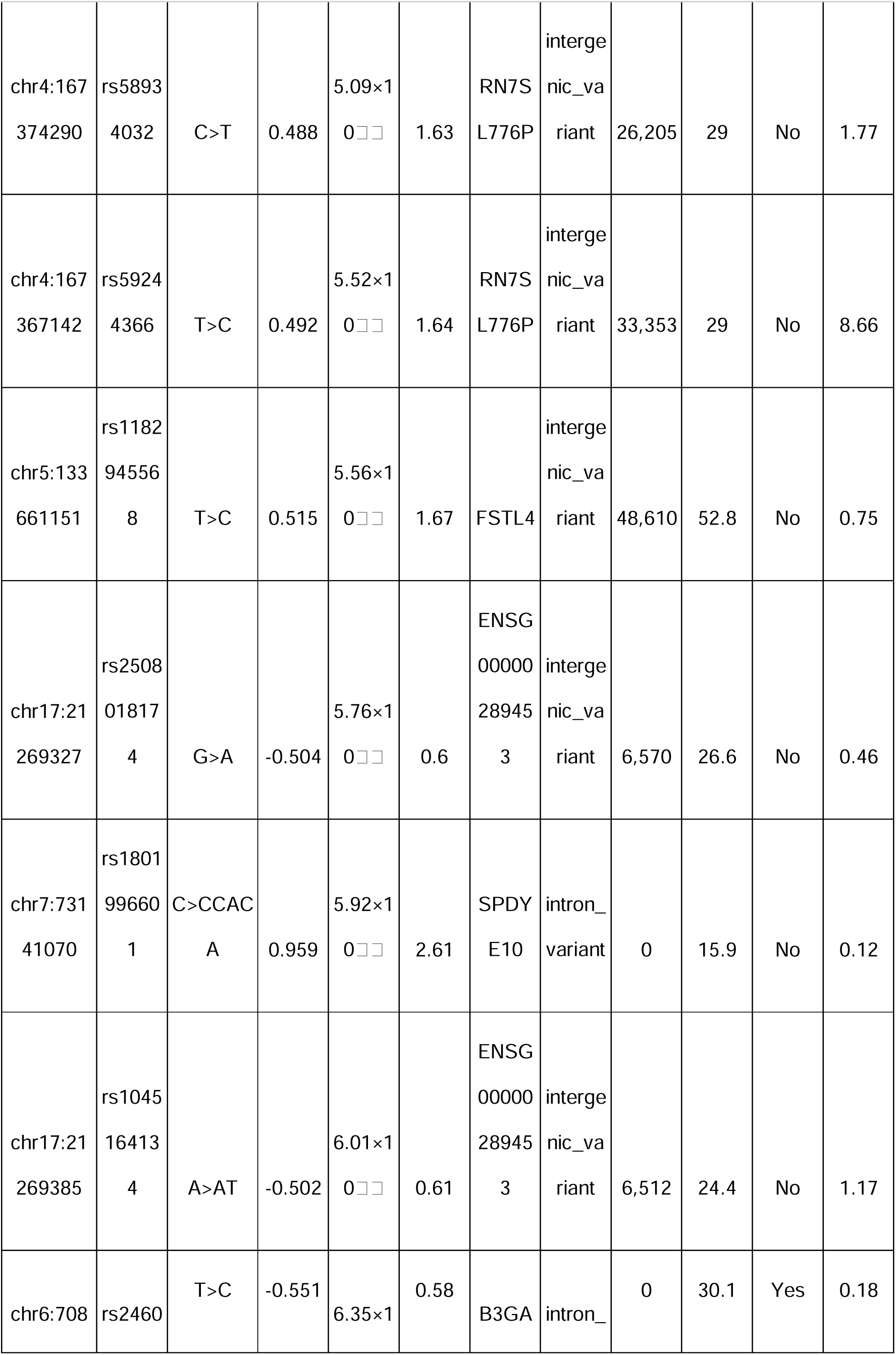
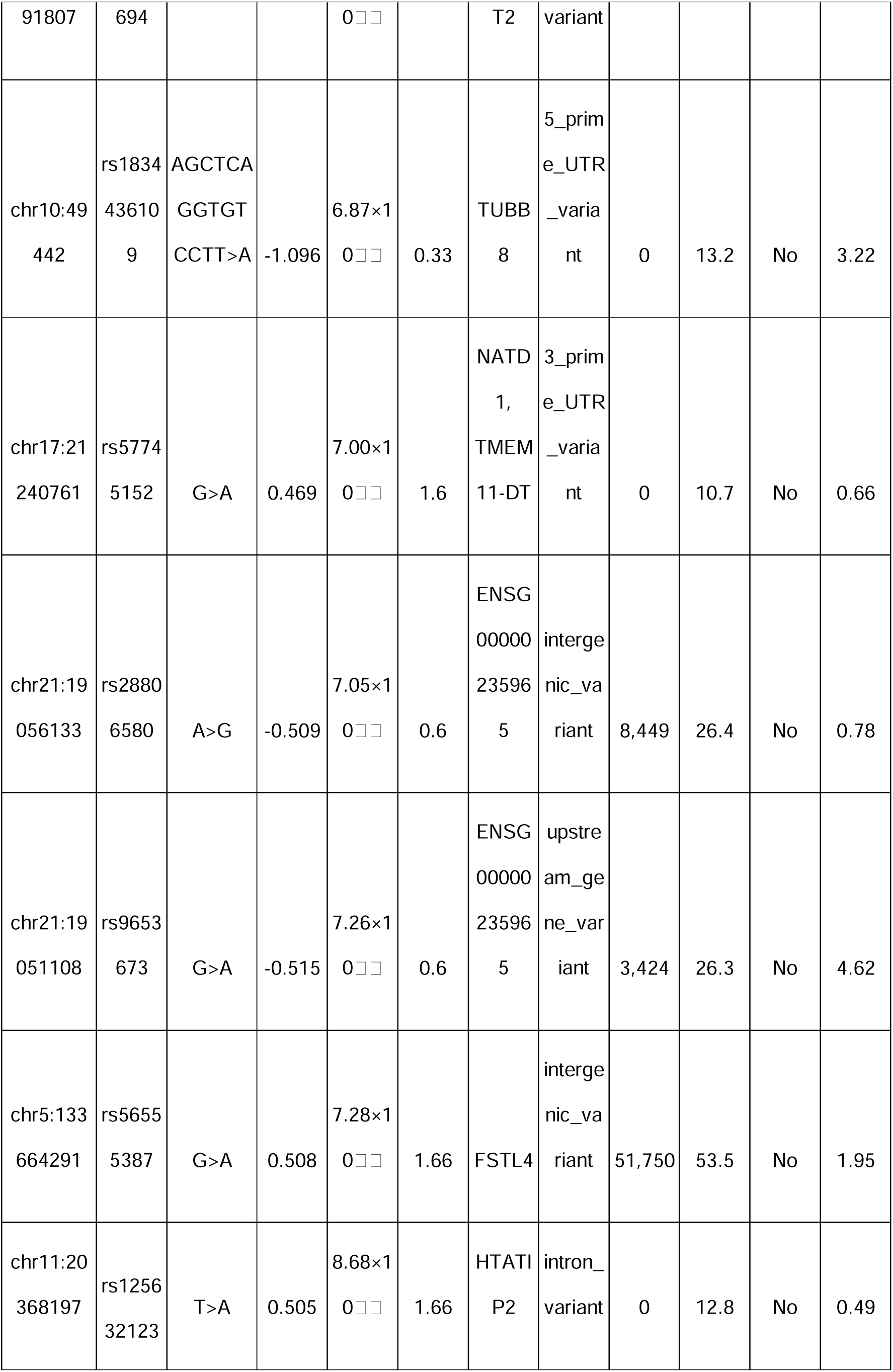
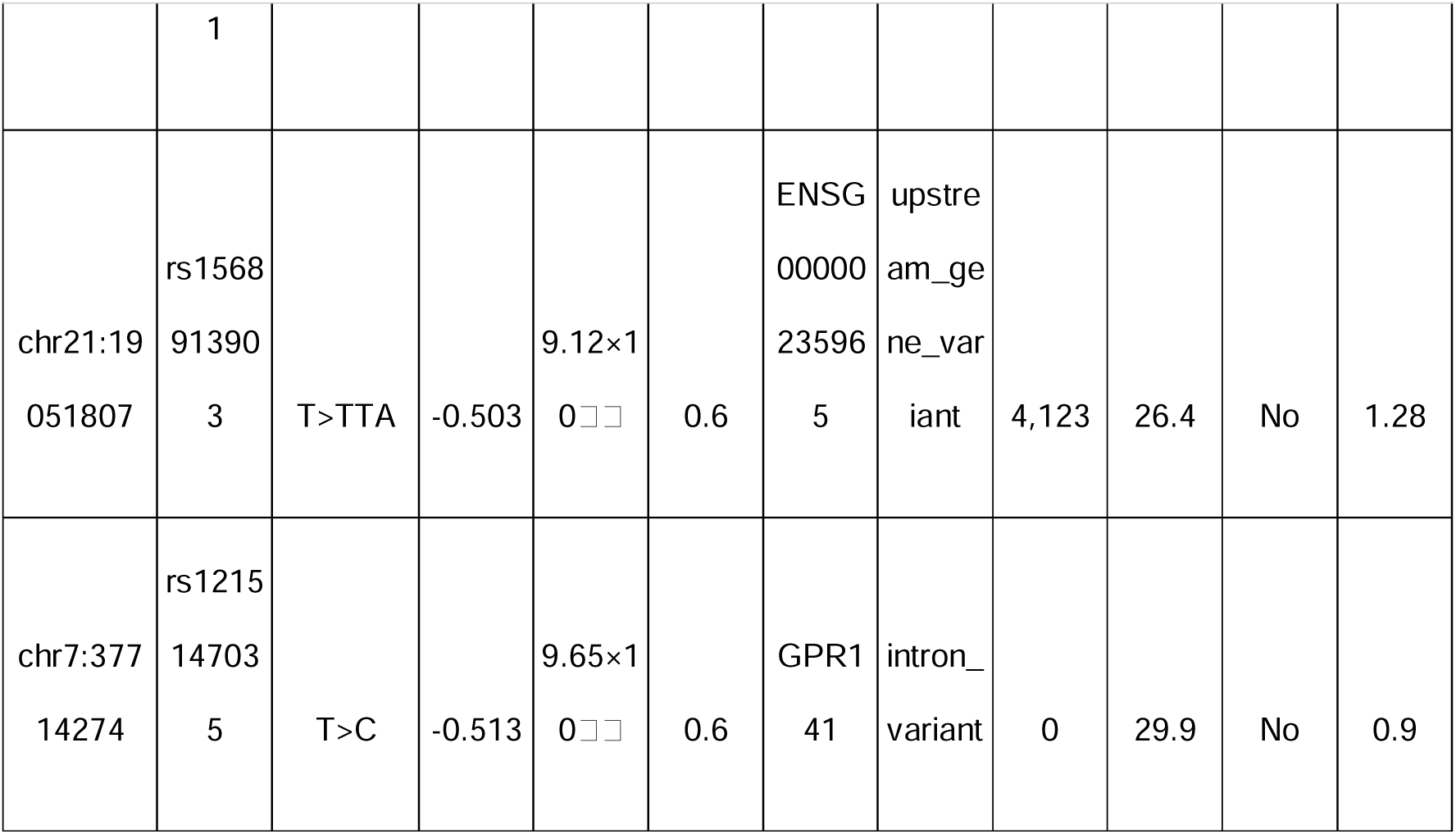
35 Suggestive SNPs from ACAF analysis.

### Functional and Regulatory Annotation of Suggestive Variants

#### Variant Characterization and Genomic Context

Comprehensive functional annotation of the 38 suggestive variants revealed a characteristic GWAS architecture dominated by non-coding variation. Variant Effect Predictor (VEP) analysis demonstrated that 94.7% of variants were non-coding, with intergenic variants comprising the largest category at 44.7% (17/38), followed by intronic variants at 34.2% (13/38), upstream gene variants at 7.9% (3/38), 3’ UTR variants at 5.3% (2/38), and one each of 5’ UTR variant, synonymous variant, and splice region variant.

For intergenic variants, nearest gene analysis revealed that all variants were located within 500kb of at least one gene, with 18 variants (47.4%) residing within gene boundaries (0 bp distance) and 20 variants (52.6%) located near genes at distances ranging from 3,424 to 51,750 bp. Notably, nine variants clustered near the pseudogene RN7SL776P on chromosome 4 (distances 26-44kb), and five variants were associated with ENSG00000235965 on chromosome 21.

#### Population Genetics and Evolutionary Context

Population frequency analysis via gnomAD v4 revealed remarkably high allele frequencies across all variants. All 38 variants (100%) were present in gnomAD, with 37/38 (97.4%) being common variants (MAF ≥ 5%). The mean allele frequency was 30.96%, with a median of 29.60%. According to ACMG guidelines, these 37 variants meet the BA1 criterion for stand-alone benign classification based on allele frequency alone.

The sole exception was rs62420977 (chr6:129165442G>A), with an allele frequency of 2.15% in exomes and 1.88% in genomes. Population stratification analysis revealed striking enrichment in the Amish population (13.71%), with particularly high frequency in Amish females (17.23%) compared to males (9.95%). This variant showed highest frequencies in Finnish (2.37%) and Non-Finnish European (2.95%) populations, while being nearly absent in East Asian (0.02%) and having low frequency in African (0.47%) populations.

#### Pathogenicity Prediction

CADD v1.7 analysis uniformly indicated benign consequences. All 38 variants had PHRED-scaled scores below 10, with a mean of 2.06 ± 2.20 and maximum of 8.66. The highest-scoring variant (chr4:167367142T>C, PHRED 8.66) remained well below the threshold of 20 typically used to identify potentially pathogenic variants. These uniformly low scores as demonstrated in Figures 5 and 6, suggest the variants are unlikely to cause disease through protein-damaging mechanisms.

**Figure 5:**
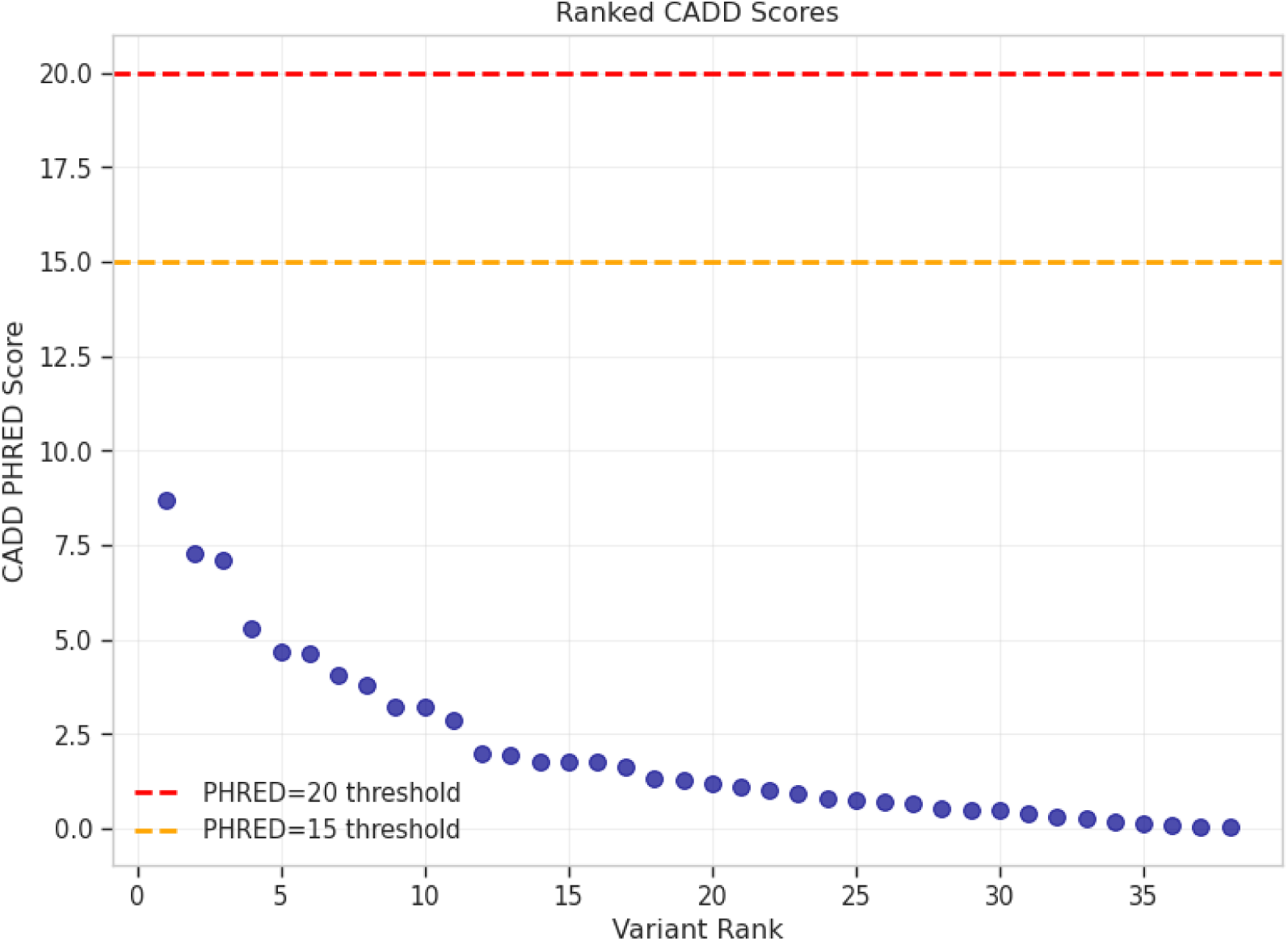
Ranked CADD PHRED scores for 38 PAD-associated variants showing all scores below pathogenicity thresholds (PHRED=15 orange line, PHRED=20 red line), with maximum score of 8.66

**Figure 6:**
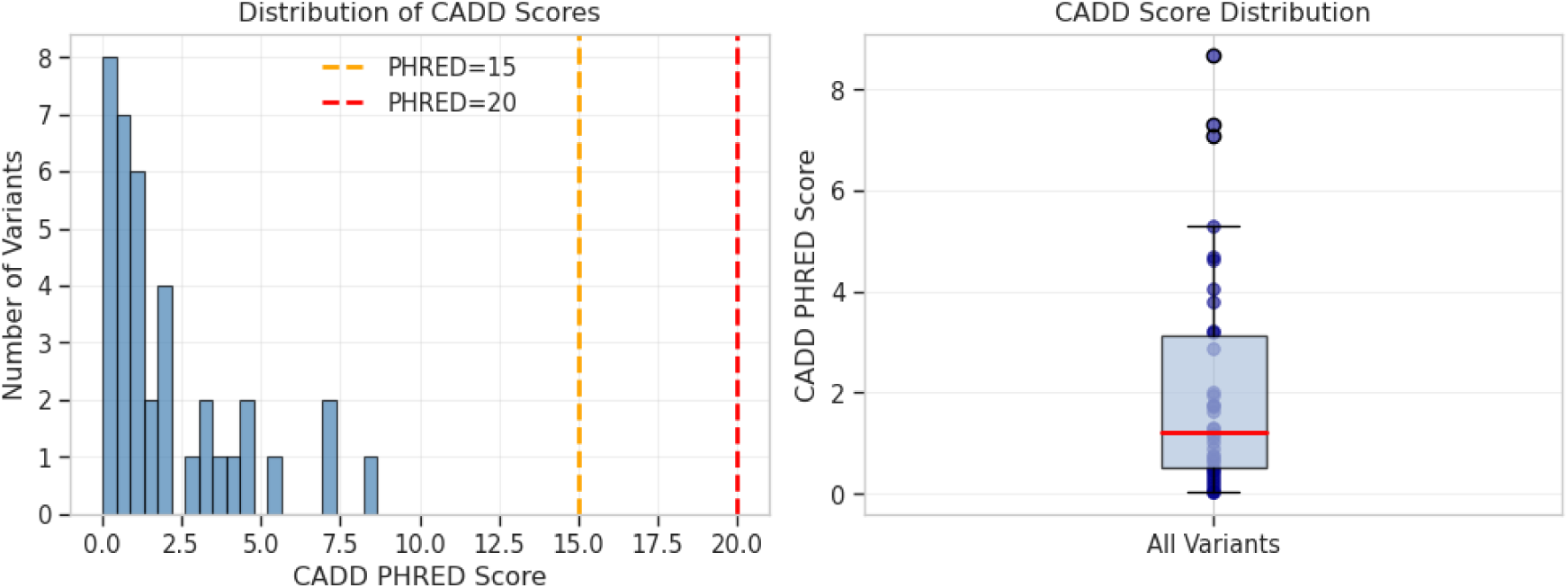
Distribution of CADD pathogenicity scores. Left: Histogram demonstrating all variants cluster below clinical significance thresholds. Right: Box plot showing median PHRED score of 1.23 (IQR: 0.50-3.12) with all variants in the benign range.

#### Splicing Analysis

The splice region variant rs267607236 in MLC1 was evaluated using SpliceAI. Despite its VEP annotation as a splice_region_variant, SpliceAI predicted no significant splicing impact, with all delta scores below 0.1 (maximum Δ = 0.06), well below the 0.2 threshold for predicted splicing effects.

#### Expression Quantitative Trait Loci Analysis

GTEx v10 analysis was limited by database coverage, with only 7/38 variants (18.4%) present in the database. Among these, three variants (7.9%) demonstrated significant quantitative trait loci effects:

1. **rs6010165** (MLC1): Significant eQTL in whole blood (p = 2.9×10□¹³, NES = -0.18), affecting expression of MLC1, MOV10L1, and PANX2. Also showed splicing QTL activity.
2. **rs62420977** (LAMA2): Splicing QTL in thyroid tissue (p = 1.8×10□□, NES = -0.56).
3. **rs2460694** (B3GAT2): eQTL in tibial nerve (p = 3.5×10□□, NES = 0.13).

Four additional variants (rs62332335, rs12108386, rs28806580, rs9653673) were present in GTEx but showed no significant eQTL associations. Notably, no variants showed eQTLs in arterial tissues (aorta, coronary, tibial arteries).

#### Regulatory Element Analysis

VEP identified only one variant, rs58661519, as overlapping a regulatory element (enhancer ENSR10_93T3ZW). However, ENCODE SCREEN analysis found no candidate cis-regulatory elements (cCREs) at any of the three positions queried (chr17:21269385, chr22:50076844, chr11:20368196) across 1,518 biosamples examined.

RegulomeDB analysis was successful for only 15/38 variants (39.5%), with two achieving score 1f (rs1045164134 and rs6010165), indicating strong regulatory evidence including eQTL data. However, the limited coverage suggests many variants occur in poorly characterized regulatory regions.

#### Chromatin Accessibility Analysis

ENCODE chromatin accessibility data was available for all variants but was limited to five non-vascular cell types: HCT116 cells, activated and resting T-cells, A673 cells, and LPS-treated macrophages. No PAD-relevant vascular tissue data was available, preventing tissue-specific interpretation.

### Comparison of Results Across Analyses

#### Unified Genetic Architecture

The integration of multiple analytical approaches revealed a remarkably consistent genetic architecture underlying PAD amputation risk. Conditional analysis across seven genomic regions containing multiple suggestive variants demonstrated uniform absence of secondary signals. These regions encompassed substantial genomic territory, with variant counts ranging from 682 in the chromosome 17 region to 1,056 in the chromosome 11 region. After conditioning on the lead variant in each region, all association signals were effectively eliminated, with conditional p-values showing dramatic increases typically of several orders of magnitude compared to the original associations.

This pattern held true even for the chromosome 4 region at 167.3-167.4Mb, which contained nine of our suggestive variants representing the highest density in our dataset. The complete elimination of secondary signals across all regions indicates that each associated locus contains a single causal variant, with all other associations merely reflecting linkage disequilibrium. This relatively simple genetic architecture contrasts with many complex traits that show extensive allelic heterogeneity and suggests that identifying causal variants through fine-mapping may be more tractable than initially anticipated.

#### Critical Impact of Study Design

The alternative control analysis provided compelling validation of our extreme phenotype design, revealing how profoundly study design choices influence genetic discovery. When we compared our matched design utilizing 399 cases and 405 controls against an analysis using all available PAD patients without amputation as controls, comprising 399 cases and 7,139 controls, the results were striking. Despite the 18-fold increase in control sample size, which traditional power calculations would suggest should enhance discovery, we observed a paradoxical and dramatic loss of statistical power. The systematic loss of signal when using all controls versus our extreme phenotype design is visualized in Figure 7, demonstrating how the traditional approach dilutes amputation-specific genetic effects.

**Figure 7:**
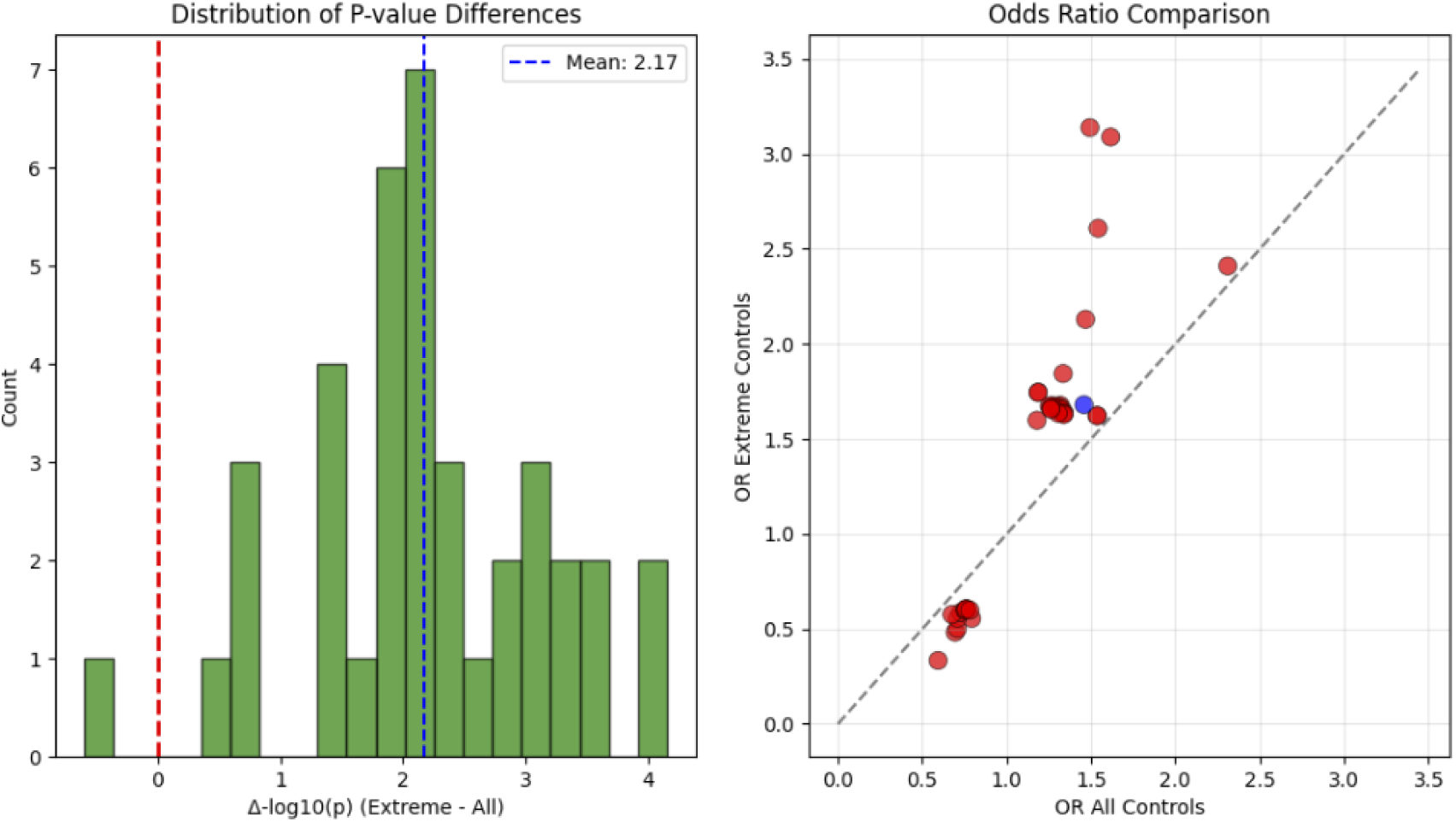
Alternative control analysis comparing extreme phenotype design versus all PAD controls. Left: Distribution of p-value differences (Δ-log□□(p)) showing 37/38 variants with improved significance using extreme phenotype design (positive values), with mean improvement of 2.17 orders of magnitude. Right: Odds ratio comparison demonstrating systematic attenuation of effect sizes when using all controls, with red points indicating variants that lost significance and the single blue point (rs60516505) showing improved association with all controls.

Of the 38 variants that achieved suggestive significance in our extreme phenotype design, 36 variants lost this significance threshold when tested with all controls, representing a 94.7% failure rate. The mean p-values increased by 2.17 orders of magnitude, approximately 148-fold, while mean odds ratios attenuated from 1.42 to 1.16, representing a 14.6% reduction in apparent effect size. Several variants showed particularly dramatic deterioration in statistical evidence. The variant rs59299975 on chromosome 11 saw its p-value deteriorate from 2.00×10□□ to 0.028, a staggering 14,000-fold increase that would lead to its complete dismissal in a conventional analysis. Similarly, rs57745152 shifted from 7.00×10□□ to 0.026, and rs1834436109 moved from 6.87×10□□ to 0.012.

Intriguingly, only one variant, rs60516505 on chromosome 19, showed modest improvement with the all-controls design, with its p-value changing from 2.54×10□□ to 6.44×10□□. This unique behavior suggests this variant may capture genetic risk for general PAD severity rather than the specific progression to amputation, highlighting how different study designs can capture distinct aspects of disease biology.

#### Robustness to Demographic Confounding

The stability of our genetic findings to demographic adjustment provided important validation of their biological relevance. Our covariate adjustment analysis successfully tested all 38 candidate variants in 859 individuals with complete phenotype and genotype data, comprising 424 amputation cases and 435 controls. When comparing models incorporating only ancestry principal components against models that additionally included age at enrollment, the results demonstrated remarkable consistency.

The p-values from the baseline and age-adjusted models showed near-perfect correlation with r = 0.999, indicating that age adjustment had minimal impact on the genetic associations. All 38 variants that showed suggestive association in the baseline model maintained this level of significance after age adjustment, with no variant losing its suggestive status. The mean - log□□(p-value) across all variants increased slightly from 4.857 in the baseline model to 4.887 in the age-adjusted model, suggesting that accounting for age, if anything, slightly strengthened rather than weakened the genetic associations. The minimal impact of age adjustment on genetic associations is shown in Figure 8, with points clustering tightly along the diagonal.

**Figure 8:**
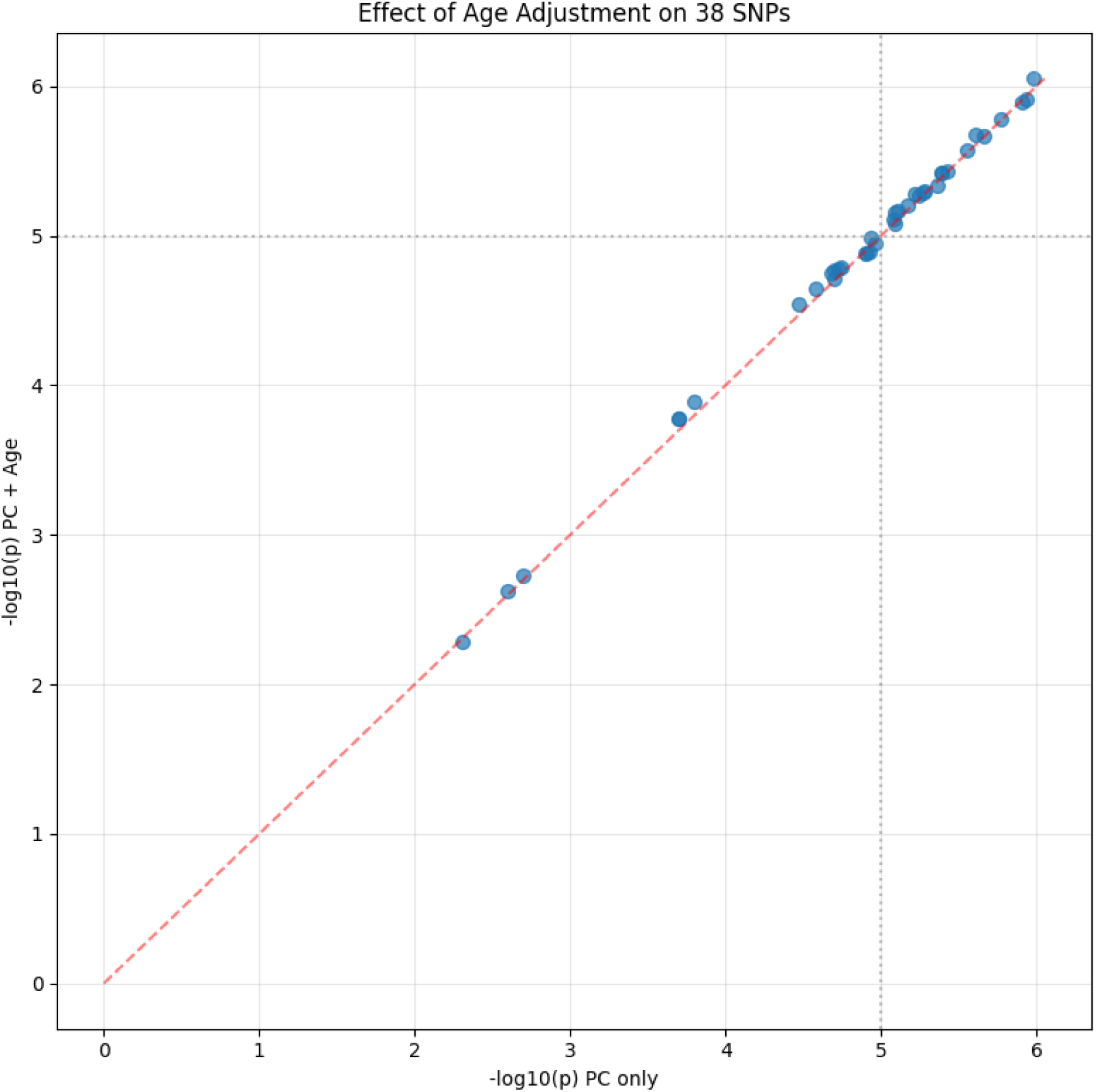
impact of age adjustment on genetic associations

Individual variants showed minimal change, with the strongest association at rs1256321231 in the HTATIP2 gene improving from p = 1.04×10□□ to p = 8.89×10□□ after age adjustment. Among the top associations, the maximum change in p-value was less than one order of magnitude, with most variants showing changes of less than 10% in their -log□□(p-value).

While we attempted to additionally adjust for sex, these models failed to converge due to numerical instability, likely reflecting the strong sex imbalance with approximately 62% male representation among amputation cases. However, given the extreme stability observed with age adjustment and the fact that ancestry principal components partially capture population-level sex differences, this limitation is unlikely to substantially impact our conclusions.

#### Functional Annotation Integration

The comparison of results across multiple functional annotation platforms revealed both the current limitations of functional genomics resources for vascular diseases and the convergent evidence supporting a small subset of our variants. Database coverage showed marked disparities that highlight the fragmented nature of current functional annotation resources.

While ENCODE provided chromatin accessibility data for all 38 variants, this apparent complete coverage was misleading as the data consistently came from the same five non-vascular cell types across all queries, including colorectal cancer cells, T-cells, Ewing sarcoma cells, and macrophages. This uniformity prevented any tissue-specific interpretation relevant to peripheral artery disease.

The Genome Aggregation Database provided complete coverage and revealed that 37 of 38 variants were common in human populations with allele frequencies exceeding 5%, definitively excluding rare variant models of disease. GTEx offered more limited coverage at only 18.4%, successfully querying just 7 of 38 variants, but yielded the most functionally relevant findings with three confirmed expression quantitative trait loci. RegulomeDB showed intermediate coverage at 39.5%, annotating 15 variants with two achieving the highest regulatory evidence scores. Most strikingly, ENCODE SCREEN failed to identify any candidate cis-regulatory elements for the specific positions we queried, despite examining 1,518 biosamples.

Despite this limited overlap across platforms, certain variants emerged with multiple lines of supporting evidence. The variant rs6010165 demonstrated convergent support as a blood eQTL with highly significant effects on gene expression, achieved a RegulomeDB score of 1f indicating comprehensive regulatory evidence, and represented a synonymous coding change that might still affect regulation. The variant rs62420977 stood out as both a splicing QTL affecting LAMA2 and the sole low-frequency variant in our set with a minor allele frequency of 2.15%, suggesting potential functional importance. The variant rs58661519 was notable for its location within an enhancer element and its intronic position within ADAM12, a metalloproteinase gene relevant to vascular remodeling.

The complete absence of arterial eQTLs across aorta, coronary, and tibial artery tissues in GTEx represents a critical gap, as these tissues would provide the most direct functional evidence for variants affecting peripheral artery disease. Similarly, the lack of vascular-specific chromatin accessibility data from ENCODE means we may be missing crucial tissue-specific regulatory mechanisms that only manifest in disease-relevant cell types.

#### Regional Architecture Patterns

The regional association analyses corroborated our conditional analysis findings while revealing interesting patterns of variant clustering. Analysis of the top six loci demonstrated that five regions showed isolated association peaks with minimal flanking signals, consistent with single causal variants driving the associations. The lead variants in these regions rose clearly above background noise with no accompanying signals approaching significance, supporting a simple genetic architecture. Regional association plots for the six most suggestive loci is shown in figure 9. Each panel shows one locus with ±500kb (or ±1Mb for chr4:167Mb region) flanking the lead SNP (purple diamond). Colors indicate approximate linkage disequilibrium based on physical distance from lead variant. Horizontal lines mark genome-wide (red) and suggestive (orange) significance thresholds.

**Figure 9:**
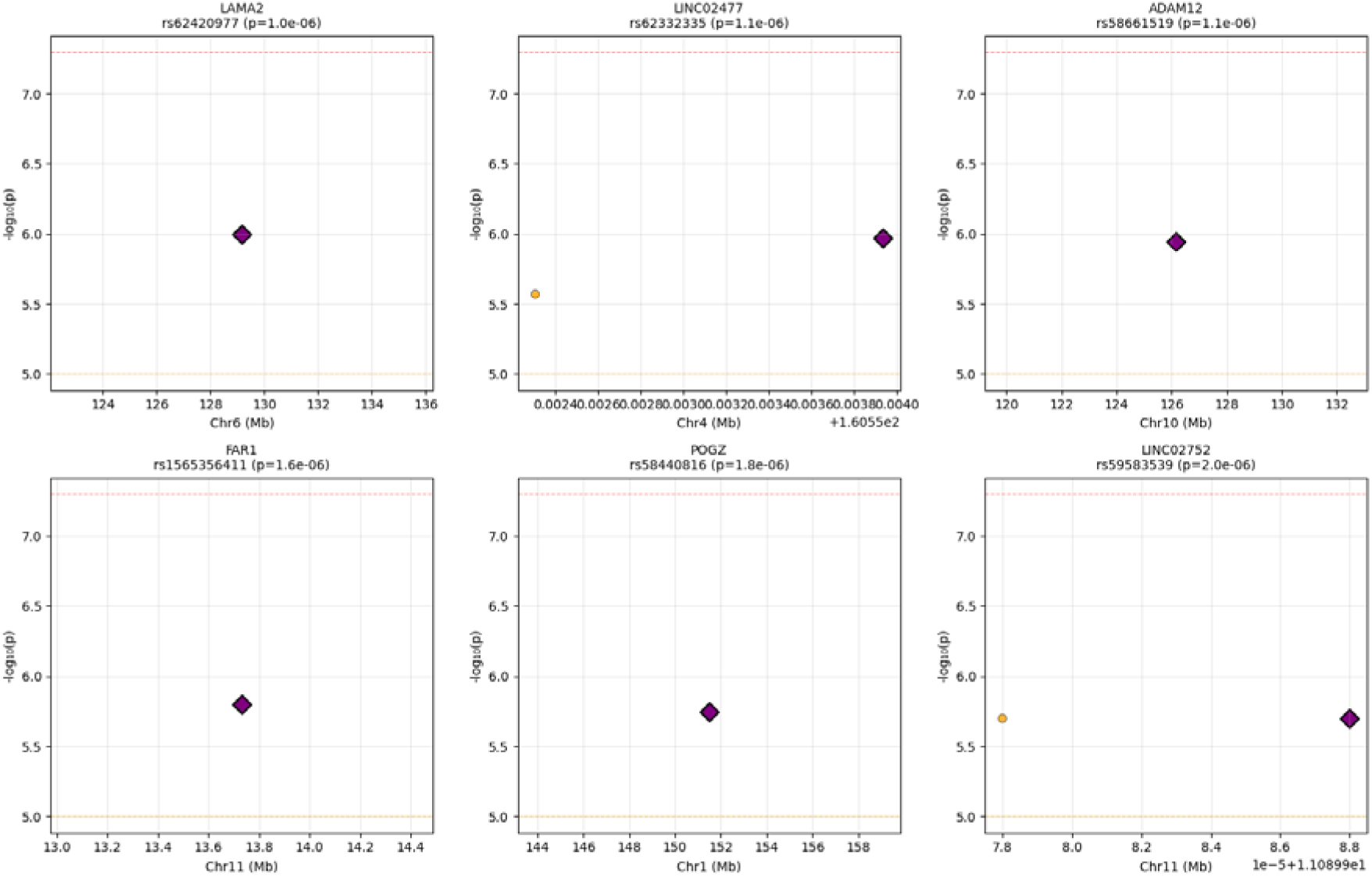
Regional association plots for the six most suggestive loci

The LINC02477 locus on chromosome 4 represented the sole exception to this pattern, displaying two variants that reached suggestive significance within approximately 1.6 kilobases of each other. These variants, rs62332335 at position 160,553,934 and rs377194319 at position 160,552,306, initially suggested possible allelic heterogeneity. Similarly, the LINC02752 locus contained two variants (rs59299975 and rs59583539) separated by only 10 base pairs with identical association statistics, likely representing the same signal captured twice or variants in perfect linkage disequilibrium. Our conditional analysis definitively demonstrated that both loci represent single association signals rather than independent effects.

The concentration of variants in certain genomic regions revealed intriguing patterns that warrant further investigation. Nine variants clustered near the pseudogene RN7SL776P on chromosome 4, with distances ranging from 26 to 44 kilobases, raising questions about whether this represents a regulatory hotspot affecting nearby genes or potentially reflects technical artifacts from repetitive sequences. Similarly, five variants associated with ENSG00000235965 on chromosome 21 suggest another potential regulatory domain. The identification of variants affecting long non-coding RNAs, specifically LINC02477 and LINC02752, aligns with growing recognition of lncRNA roles in vascular biology and disease, though their specific functions remain largely uncharacterized.

## Discussion

This study represents the first genome-wide investigation specifically targeting genetic determinants of amputation risk in PAD patients. Our approach, employing both targeted GWAS using Clinvar analysis and comprehensive GWAS using ACAF methodologies, has identified several suggestive genetic loci that may contribute to amputation risk in this population.

### Interpretation of Key Findings

This first genome-wide investigation of genetic determinants specific to amputation risk in PAD patients identified 38 suggestive loci, though none achieved genome-wide significance. The absence of genome-wide significant findings (p < 5×10□□) with our limited sample size likely reflects limited statistical power rather than absence of genetic effects, given the observed effect sizes ranging from OR 0.48 to 3.14.

The predominance of non-coding variants at 94.7% aligns with the established GWAS paradigm that regulatory variation, rather than protein-coding changes, drives most complex trait associations. Among our 38 variants, only five showed potential for direct functional impact: one splice region variant (rs267607236 in MLC1), 1 synonymous variant (rs6010165), two 3’ UTR variants (rs1565356411 in FAR1 and rs57745152 in NATD1), and one 5’ UTR variant (rs1834436109 in TUBB8). However, computational predictions suggested limited functional consequences, with the MLC1 splice variant showing SpliceAI scores below significance thresholds and all variants having CADD scores below 10.

The functional evidence was strongest for three variants confirmed as expression quantitative trait loci. The rs6010165 variant in MLC1 demonstrated a highly significant blood eQTL (p = 2.9×10□¹³) affecting multiple genes including MLC1, MOV10L1, and PANX2, highlighting potential systemic inflammatory mechanisms in amputation risk. The rs62420977 variant in LAMA2, encoding a critical vascular basement membrane component, showed splicing QTL activity that could affect protein isoforms important for vascular integrity. Additionally, rs2460694 in B3GAT2 showed a weak eQTL signal in GTEx Nerve - Tibial tissue (p = 3.5×10□□, NES = 0.13).

Several genes implicated by our analysis have plausible biological roles in PAD progression. ADAM12 encodes a metalloproteinase involved in extracellular matrix remodeling, CUBN is involved in vitamin B12 absorption potentially affecting homocysteine levels, and FAR1 catalyzes fatty alcohol synthesis relevant to lipid metabolism. The identification of protective variants, such as rs58440816 near POGZ with OR=0.56, suggests some genetic factors may confer resilience against progression to amputation.

The remarkably high population frequencies, with 37 of 38 variants showing MAF above 5% and a mean frequency of 31%, definitively exclude Mendelian inheritance patterns. This frequency distribution, combined with uniformly low CADD scores, supports a polygenic architecture where multiple common variants with individually small effects collectively influence amputation risk.

### Methodological Considerations

Our extreme phenotype design comparing amputation cases to matched PAD controls proved crucial for discovery. The alternative analysis demonstrated that using all 7,139 available PAD patients as controls, despite providing 18-fold more samples, would have missed 94.7% of our associations. This paradox occurs because including mild disease cases dilutes genetic signals specific to severe outcomes. Mean p-values deteriorated by 2.17 orders of magnitude in the conventional analysis, with some variants showing 14,000-fold increases in p-values.

The uniform absence of secondary signals in conditional analysis across all seven tested regions simplifies interpretation and future fine-mapping efforts. Each region appears to harbor a single causal variant, with other associations reflecting linkage disequilibrium. This contrasts with many complex traits showing multiple independent signals per locus.

The remarkable stability of associations with age adjustment, showing correlation of 0.999 between models, indicates these genetic factors operate independently of chronological age. This is particularly noteworthy given age is a major clinical risk factor for amputation, suggesting these variants influence biological pathways rather than simply correlating with disease duration.

Several technical constraints affected our analyses. Models adjusting for sex failed to converge due to the imbalanced sex distribution. The cross-sectional design prevented analysis of genetic effects on time to amputation. Most critically, functional annotation was severely limited by the scarcity of vascular-specific data in public databases, with no arterial eQTLs found in GTEx despite the vascular nature of PAD, and ENCODE chromatin accessibility data lacking vascular cell types for our queried regions.

### Biological and Clinical Implications

Our findings support a model where amputation risk results from modest perturbations across multiple biological systems rather than severe disruption of single pathways. The two strongest eQTLs suggest mechanisms involving systemic inflammation (MLC1 blood eQTL, p=2.9×10□¹³) and vascular structural integrity (LAMA2 splicing QTL, p=1.8×10□□), while a nominal association with B3GAT2 in nerve tissue (p=3.5×10□□) suggests possible peripheral nerve involvement. However, the limited functional evidence for most variants indicates that current genomic databases inadequately capture vascular-specific regulatory mechanisms.

The high population frequencies of associated variants suggest they are well-tolerated under normal conditions but may lower the threshold for irreversible ischemic damage when combined with environmental stressors and traditional risk factors. The mix of protective and risk variants, with odds ratios ranging from 0.48 to 3.14, indicates complex balancing of genetic factors influencing progression to amputation.

For clinical translation, these findings suggest polygenic risk scores rather than single variant testing will be needed for risk stratification. However, the current variants explain limited phenotypic variance and would require validation before clinical implementation. The population-specific enrichment of rs62420977 in Amish populations (13.7% vs 2.15% in gnomAD exomes) highlights the need for ancestry-aware approaches in precision medicine.

### Comparison with Previous Literature

As the first genome-wide association study specifically examining genetic determinants of amputation risk in peripheral artery disease patients, our findings lack direct comparisons in the existing literature. Previous genetic studies of PAD have focused primarily on disease susceptibility rather than progression to severe outcomes such as limb loss.

The most relevant prior work includes the PAD GWAS by Klarin et al. (2019) in the Million Veteran Program, which identified 19 loci associated with PAD susceptibility in over 31,000 cases. However, their phenotype definition included all PAD patients without stratification by amputation status. Similarly, Matsukura et al. (2015) conducted a GWAS in a Japanese population identifying susceptibility loci for PAD, but did not examine amputation as an outcome. Our approach of comparing amputation cases to matched PAD controls without amputation represents a methodological advance in studying genetic factors specific to disease progression rather than initiation.

The absence of overlap between our suggestive loci and previously reported PAD susceptibility variants supports the hypothesis that genetic architecture underlying disease complications differs from that influencing disease susceptibility. This aligns with observations in other complex diseases where progression genetics show distinct patterns from susceptibility genetics.

Our findings of predominantly common variants with modest effect sizes (OR 0.48-3.14) parallel the genetic architecture observed in other cardiovascular traits. The high population frequencies we observed (mean MAF 31%) are consistent with the polygenic model of complex disease, similar to findings in coronary artery disease GWAS where most associated variants are common with small individual effects.

The lack of genome-wide significant findings with our sample size mirrors early GWAS efforts in other vascular diseases. Initial PAD GWAS studies similarly required meta-analyses of multiple cohorts to achieve genome-wide significance, suggesting our sample of 804 individuals represents an important first step requiring expansion through collaborative efforts.

### Strengths and Limitations

This study’s primary strength lies in its novel phenotype definition specifically targeting amputation risk rather than PAD susceptibility. The extreme phenotype design maximized statistical power from a moderate sample size by comparing phenotypic extremes.

Comprehensive multi-platform functional annotation, though revealing limited functional evidence, provided a thorough characterization attempt. The All of Us cohort offers greater ancestral diversity than traditional European-focused studies. Multiple sensitivity analyses, including conditional analysis and alternative control comparisons, confirmed the robustness of our approach.

However, significant limitations must be acknowledged. The sample size of 804 individuals limited power to achieve genome-wide significance or detect variants with smaller effects. No independent replication cohort was used to validate findings. Functional annotation was severely constrained by the absence of vascular-specific data in public databases, with no arterial eQTLs or relevant chromatin accessibility data. We could not adjust for important clinical variables including diabetes, smoking, or medications. The cross-sectional design prevented temporal analysis of progression. Additionally, database coverage was incomplete, with only 18.4% of variants in GTEx and 39.5% in RegulomeDB.

## Future Directions

These findings establish a foundation for larger collaborative efforts needed to achieve genome-wide significance and enable robust replication. Functional validation of the two eQTL variants (rs6010165 and rs62420977) in disease-relevant cell types - including vascular cells for LAMA2 and blood/immune cells for MLC1 - represents an immediate priority Development of vascular-specific functional genomics resources, including arterial eQTL datasets and endothelial cell chromatin accessibility maps, is critical for understanding these associations. Longitudinal studies tracking progression from PAD to amputation would clarify temporal relationships between genetic factors and clinical outcomes. Integration with clinical risk factors in predictive models could assess the added value of genetic information for patient stratification.

## Conclusion

This comprehensive genome-wide association study represents the first systematic investigation of genetic factors specifically influencing amputation risk in peripheral artery disease patients. Through careful extreme phenotype design, we identified 38 suggestive genetic associations that implicate diverse biological pathways in disease progression. While these variants did not achieve genome-wide significance and show limited functional annotation with current resources, the study design successfully isolated genetic factors specific to amputation risk rather than general PAD susceptibility. The simple genetic architecture with single variants per locus and the age-independent nature of associations provide optimism for future fine-mapping efforts. These results establish that progression to amputation involves genetic factors distinct from disease susceptibility, supporting precision medicine approaches that consider disease stage. Larger collaborative studies, functional validation, and development of vascular-specific genomic resources represent critical next steps in translating these findings to improved clinical outcomes for PAD patients at risk for limb loss.

## Acknowledgments

We gratefully acknowledge *All of Us* participants for their contributions, without whom this research would not have been possible. We also thank the National Institute of Health’s *All of Us* Research Program for making available the participant data [and/or samples and/or cohort] examined in this study.

## Data Availability Statement

The linked genotype and phenotype data analyzed in this study are from the National Institutes of Health’s *All of Us* Research Program and are available through its controlled-access Researcher Workbench. Due to the terms of the Data Use Agreement designed to protect participant privacy and data security, the raw and individual-level data cannot be downloaded or deposited into external public repositories. The full summary-level statistics from our genome-wide association study are available upon reasonable request to the corresponding author. All qualified researchers can apply for access to the same raw dataset by registering and completing the required ethics training via the *All of Us* Research Hub at https://www.researchallofus.org/.

